# Neutrophil-to-lymphocyte ratio on admission to predict the severity and mortality of COVID-19 patients: a meta-analysis

**DOI:** 10.1101/2020.09.14.20191098

**Authors:** Daniel Martin Simadibrata, Julius Calvin, Alya Darin Wijaya, Naufal Arkan Abiyyu Ibrahim

## Abstract

The neutrophil-to-lymphocyte ratio (NLR), an inflammatory marker, was suggested to be predictive of severity and mortality in COVID-19 patients, thus allowing early risk stratification. In this study, we investigated whether NLR levels on admission could predict the severity and mortality of COVID-19 patients. A literature search was conducted on 23 July 2020 to retrieve all published articles, including grey literature and preprints, investigating the association between on-admission NLR values and severity or mortality in COVID-19 patients. The risk of bias was assessed using the Newcastle Ottawa Scale (NOS). A meta-analysis was performed to determine the overall standardized mean difference (SMD) in NLR values and the pooled risk ratio (RR) for severity and mortality with the 95% Confidence Interval (95%CI). Meta-regression analysis was done to identify potential confounders. A total of 38 articles, including 5699 patients with severity outcomes and 6033 patients with mortality outcomes, were included. The meta-analysis showed that severe and non-survivors of COVID-19 had higher on-admission NLR levels than non-severe and survivors (SMD 0.88; 95%CI 0.72-1.04; I^2^=75% and 1.68; 95%CI 0.98-2.39; I^2^=99%, respectively). Regardless of the different NLR cut-off values, the pooled mortality RR in patients with elevated vs. normal NLR levels was 2.75 (95%CI 0.97-7.72). Meta-regression analysis showed that the association between NLR levels on admission and COVID-19 severity and mortality was unaffected by age (p=0.236; p=0.213, respectively). High NLR levels on admission were associated with severe COVID-19 and mortality. Further studies need to focus on determining the optimal cut-off value for NLR before clinical use.

**Key Points:**

- High neutrophil-to-lymphocyte ratio (NLR) is associated with severe COVID-19 and mortality
- NLR is a simple, accessible, near real-time, and cost-effective biomarker recommended for use in resource-limited healthcare settings

## Introduction

As of 11 March 2020, over 120 000 Coronavirus disease 2019 (COVID-19) cases were confirmed globally, resulting in its declaration as a pandemic^1^. COVID-19 is caused by the Severe Acute Respiratory Syndrome Coronavirus-2 (SARS-CoV-2), a novel virus in the same cluster as the SARS-CoV-1 and MERS-CoV, that previously caused outbreaks in 2003 and 2012^2,3^. The clinical manifestations of COVID-19 patients range from asymptomatic to severe symptoms. A minority (30%) progresses into severe manifestations such as acute respiratory distress syndrome (ARDS), severe pneumonia, septic shock, coagulopathy, and death^4^. This rapid progression to severe conditions is caused by an overwhelming inflammation, known as the cytokine storm.

Biomarkers allowing prediction of disease severity in COVID-19 are urgently needed to address the problem of resource scarcity in this pandemic^5^. Early risk stratification for COVID-19 patients upon hospital admission is the key to providing optimal interventions and to carefully allocate the ongoing scarce human and technical resources^6^. This would ensure that the limited available resources are given to the right patients. The neutrophil-to-lymphocyte ratio (NLR) is an inflammatory marker derived from combining the absolute blood neutrophil and lymphocyte counts, two routinely performed parameters in clinical settings. Recently, studies have reported that NLR levels were higher in more severe patients and were suggested to confer a prognostic value in COVID-19 patients^7,8^. The underlying pathophysiology that justifies for the clinical use of this biomarker is that severe COVID-19 patients were more likely to present with higher levels of inflammation upon hospital admission. Therefore, obtaining NLR levels on hospital admission could allow early risk stratification, identifying patients who should be prioritized for scarce resources.

We performed a systematic review and meta-analysis to investigate whether clinical outcomes of severity and mortality in COVID-19 patients can be predicted with on-admission NLR values.

## Methods

This systematic review and meta-analysis is reported according to the Preferred Reporting Items for Systematic Review and Meta-analysis (PRISMA) Checklist (**Table S1**). Before writing this review, a detailed protocol was created and registered to the International Prospective Register of Systematic Reviews (PROSPERO) on 1 June 2020 (CRD42020189218).

### Eligibility Criteria

We included all research papers investigating adult patients (older than 18 years old) with COVID-19 (diagnosed with RT-PCR) that contain information on the NLR value at the time of hospital admission and clinical grouping of outcomes with a clinically validated definition of COVID-19 severity or death. The following articles were excluded from this review: non-research letters, correspondences, case reports, review articles, and original articles with samples below 20. Due to the limited resources, we only included articles published in English.

### Search Strategy

We searched for all published articles, including preprints and grey literature, from electronic databases on 23 July 2020. Peer-reviewed papers were sought from four databases (Ovid MEDLINE, Ovid EMBASE, SCOPUS, and the Cochrane Library); preprints were searched from three databases (MedRxiv, BioRxiv, and SSRN); and grey literature was searched from two databases (WHO COVID-19 Global Research Database, and the Centers for Disease Control and Prevention COVID-19 Research Article Database). The search strategies used were developed from the following key concepts: “COVID-19”, “Neutrophil-to-lymphocyte ratio”, “Severity”, and “Mortality” (**Table S2**). Manual hand-searching and forward and backward tracing of citations from relevant articles were also done to identify additional studies. All articles retrieved from the systematic searches of electronic databases were exported to EndNote X9 bibliographic and reference manager. Following deduplication, the titles and abstracts were screened independently by three reviewers (**DMS, ADW, NAAI**), and the remaining articles were screened for its full text against the eligibility criteria. Any disagreements were resolved through discussion until a common consensus was reached.

### Quality Assessment

The studies were critically appraised using the Newcastle Ottawa Scale (NOS) by three independent reviewers (**DMS, ADW, NAAI**), and when there is a discrepancy in the assessment score, discussions were done to reach an agreement.

### Data Extraction and Synthesis

Prior to the data extraction, a customized, standardized data extraction form was developed. The data extracted included: first author, year of study, publication type, study location, study design, baseline population characteristics (including age, gender, and underlying diseases such as diabetes mellitus, hypertension, and cardiovascular diseases), exposures, and outcomes. The exposure was defined as the NLR value on admission to the hospital, presented as either continuous or dichotomized NLR values. The outcomes of interest were severe COVID-19 and mortality. Severe COVID-19 was defined as patients who met any of the following features: (1) respiratory rate >30 breathes per min; (2) oxygen saturation <93% (ambient air); (3) ratio of the partial pressure of arterial blood oxygen (PaO2) / oxygen concentration (FiO2) ≤300mmHg^9^. Due to different severity grouping criteria among studies, non-severe COVID-19 included patients with either mild, moderate, common, ordinary, or any combination. Meanwhile, severe COVID-19 included patients in severe, critical, or a combination of the two. Additionally, for studies that performed and reported receiver operating characteristic (ROC) analysis on either severity or mortality, we extracted the optimal NLR cut-off values, the area under the curve (AUC), sensitivity, and specificity.

### Statistical Analysis

The quantitative data were exported to Review Manager 5.3 (Cochrane Collaboration) and Stata version 16, and a meta-analysis of studies was performed. For non-normal data, we extrapolated the mean and standard deviation from the available median and interquartile range (IQR) using the method by Hozo *et al*.^10^. For studies that reported the means of NLR among groups, pooled standardized mean difference (SMD) and the 95% Confidence Interval (95%CI) were obtained using the inverse variance method. For NLR values reported as dichotomized variables, the pooled risk ratio (RR) with the 95%CI was obtained using the Mantel-Haenszel method. Statistical heterogeneity was determined using the Cochrane chi-square and I^2^ with cut-off values for I^2^ of greater than 50% to be considered significantly heterogeneous. In this study, we used the random-effects model if I^2^ was greater than 50%, and the fixed-effects model if I^2^ was below or equal to 50%. Sensitivity analysis was done by omitting one study at a time to identify any source of heterogeneity and restricting the studies to only peer-reviewed papers and only studies with low risk of bias. Publication bias was assessed qualitatively using the funnel plot by comparing the SMD and the standard error of the natural log of SMD [SE(SMD)], and quantitatively using Egger’s linear regression test to evaluate the presence of small-study effects. A random-effects meta-regression was performed for the following potential confounders: age, gender, diabetes mellitus, hypertension, and cardiovascular disease. A statistically significant difference was considered if a two-tailed p<0.05.

## Results

### Search Selection

A total of 203 papers were identified from the peer-reviewed databases, and an additional nine papers were retrieved from manual hand-searching, preprint, and grey literature databases. After removing duplicates, 102 unique articles were reviewed for its titles and abstracts, leaving a total of 55 articles eligible for full-text review. After a thorough evaluation, according to the eligibility criteria, 38 papers met the inclusion criteria (**Figure 1**).

**Figure 1.**
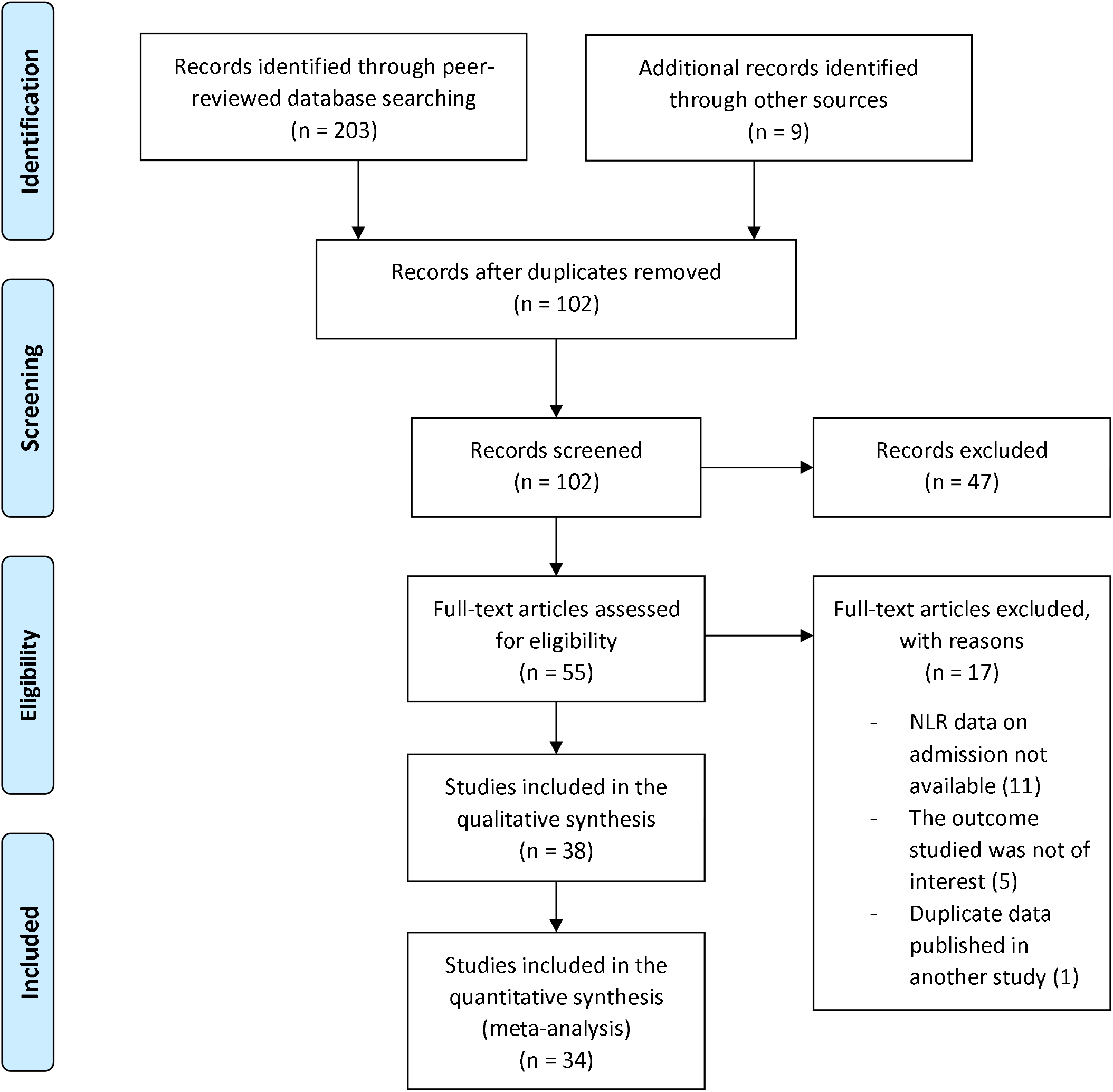
Prisma diagram for study selection. A systematic literature search was done on 23 July 2020 to identify peer-reviewed papers, preprints, and grey literature.

### Characteristics of studies

This systematic review included 38 articles incorporating 5699 patients with severity outcomes and 6033 patients with mortality outcomes. Thirty-two articles were peer-reviewed^8,11-41^, and six were preprints^42-47^; 23 articles compared NLR values on admission in severe vs. non-severe COVID-19 patients, 13 articles compared NLR values on admission in survivors vs. non-survivors of COVID-19, and only two articles compared the NLR values on admission in regard to both the severity and mortality of COVID-19 patients^20,46^. All the studies were retrospective observational studies, except for one which was prospective^32^. Most of the studies were conducted in China, with only four studies (11%) performed outside of China, among which two studies were in Turkey^39,40^, and two were in the United States of America^32,33^ (**Table 1, Table 2, Table S3-4**). Studies with severity as the outcome measure had a median risk of bias score of 7 (IQR 5.5-8.5; range 5-9). On the other hand, studies comparing the NLR value on admission in survivors and non-survivors of COVID-19 had a median risk of bias score of 8 (IQR 7-9; range 6-9) (**Table S5**).

**Table 1.** Baseline characteristics of included studies comparing severe and non-severe COVID-19 patients.

| First Author | Study Location | Groups | Sample (N) | Male % | Age (years)<br>Mean ± SD /<br>Median (IQR) | DM<br>N (%) | HT<br>N (%) | CVD<br>N (%) | NLR Value<br>Mean ± SD /<br>Median (IQR) | NOS Score |
| --- | --- | --- | --- | --- | --- | --- | --- | --- | --- | --- |
| Qin C et al | China | Severe | 286 | 54 | 61 (51-69) | 53 (19) | 105 (37) | 24 (8) | 5.5 (3.3–10.0) | 5 |
|  |  | Non-severe | 166 | 48 | 53 (41-62) | 22 (13) | 30 (18) | 3 (2) | 3.2 (1.8–4.9) |  |
| Zhang Y et al | China | Severe | 31 | 65 | 65 ± 13 | NR | NR | NR | 7.58 ± 7.04 | 7 |
|  |  | Mild | 84 | 35 | 44 ± 15 | NR | NR | NR | 2.28 ± 1.29 |  |
| Yang AP et al | China | Severe | 24 | 75 | 58 ± 12 | 13 (54) | 16 (67) | 9 (38) | 20.7 ± 24.1 | 7 |
|  |  | Non-severe | 69 | 55 | 42 ± 19 | 8 (12) | 7 (10) | 4 (6) | 4.8 ± 3.5 |  |
| Gong J et al | China | Severe | 28 | 57 | 64 (55-72) | NR | NR | NR | 3.7 (2.0-6.7) | 7 |
|  |  | Non-severe | 161 | 45 | 45 (33-62) | NR | NR | NR | 1.9 (1.4-2.9) |  |
| Zhu Z et al | China | Severe | 16 | 56 | 58 ± 12 | 0 (0) | 8 (50) | 2 (13) | 4.24 (3.00-10.87) | 7 |
|  |  | Non-severe | 111 | 66 | 50 ± 16 | 10 (9) | 23 (21) | 4 (4) | 2.75 (1.90-3.95) |  |
| Sun S et al | China | Severe | 27 | 67 | 62 (53-71) | NR | NR | NR | 8.71 (3.77-14.4) | 5 |
|  |  | Common | 89 | 47 | 47 (37-55) | NR | NR | NR | 2.41 (1.73-3.47) |  |
| Liu F et al | China | Severe | 19 | 79 | 63 (40-66) | 3 (16) | 6 (32) | 1 (5) | 3.4 (2.8-5.8) | 8 |
|  |  | Non-severe | 115 | 42 | 50 (36-64) | 7 (6) | 21 (18) | 4 (4) | 2.7 (1.8-3.7) |  |
| Fu J et al | China | Severe | 16 | 63 | 52 ± 13 | 4 (5) | 7 (9) | NR | 6.29 ± 3.72 | 6 |
|  |  | Mild/moderate | 59 | 59 | 45 ± 14 |  |  |  | 2.33 ± 1.22 |  |
| Ding X et al | China | Severe | 15 | 60 | 67 (55-76) | 5 (7) | 9 (13) | 6 (8) | 3.6 (2.4-9.6) | 7 |
|  |  | Non-severe | 57 | 42 | 46 (35-60) |  |  |  | 1.9 (1.3-2.9) |  |
| Chen R et al | China | Critical | 48 | 79 | 61 ± 14 | 5 (10) | 23 (48) | 7 (15) | 16.06 (11.26-26.35) | 9 |
|  |  | Severe | 155 | 60 | 61 ± 14 | 23 (15) | 52 (34) | 14 (9) | 8.96 (4.62-17.04) |  |
|  |  | Mild/moderate | 345 | 53 | 67 ± 12 | 33 (10) | 73 (21) | 14 (4) | 3.37 (2.05-6.65) |  |
| Shang W et al | China | Severe | 139 | 59 | 64 (54-73) | 20 (14) | 45 (32) | 25 (18) | 4.75 (2.51-9.42) | 7 |
|  |  | Non-severe | 304 | 45 | 58 (47-67) | 43 (14) | 86 (28) | 19 (6) | 2.38 (1.57-3.72) |  |
| Xie G et al | China | Severe | 12 | 83 | 52 (35-66) | 2 (17) | 4 (33) | 2 (17) | 3.0 (1.56-6.55) | 5 |
|  |  | Non-severe | 85 | 51 | 45 (32-60) | 3 (4) | 16 (19) | 5 (6) | 2.74 (2.03-3.96) |  |
| Xie L et al | China | Severe | 51 | 57 | NR | 8 (16) | 12 (24) | 6 (12) | 7.90 ± 10.20 | 5 |
|  |  | Non-severe | 322 | 52 | NR | 21 (7) | 59 (18) | 12 (4) | 2.93 ± 1.80 |  |
| Zhou Y et al | China | Moderate | 140 | 39 | 56 ± 14 | NR | NR | NR | 3.1 ± 2.41 | 5 |
|  |  | Severe | 123 | 47 | 64 ± 14 | NR | NR | NR | 11.66 ± 27.66 |  |
|  |  | Critically severe | 41 | 61 | 65 ± 13 | NR | NR | NR |  |  |
| Wu S et al | China | Severe or critical | 67 | 67 | 66 (54-73) | 8 (12) | 22 (33) | 6 (9) | 5.8 (3.3-13.0) | 7 |
|  |  | Moderate | 203 | 42 | 61 (50-68) | 27 (13) | 59 (29) | 5 (3) | 2.2 (1.5-3.4) |  |
| Kong M et al | China | Severe | 87 | 52 | 68 ± 12 | 18 (21) | 47 (54) | 11 (13) | 6.6 (2.1-11.1) | 7 |
|  |  | Mild | 123 | 48 | 53 ± 16 | 9 (7) | 32 (26) | 9 (7) | 3.3 (1.0-3.4) |  |
| Wang F et al | China | Severe | 70 | 64 | 60 (49-64) | NR | NR | NR | 2.72 (1.87-4.37) | 8 |
|  |  | Non-severe | 253 | 43 | 41 (32-56) | NR | NR | NR | 1.72 (1.19-2.53) |  |
| Liao D et al | China | Critical | 86 | 71 | 68 (61-78) | 17 (20) | 28 (33) | 8 (9) | 16.02 (6.49-24.79) | 7 |
|  |  | Severe | 145 | 52 | 67 (58-76) | 30 (21) | 49 (34) | 8 (6) | 4.71 (2.62-7.78) |  |
|  |  | Moderate | 149 | 46 | 56 (42-68) | 14 (9) | 37 (25) | 4 (3) | 2.67 (1.69-4.08) |  |
| Ok F et al | Turkey | Severe | 54 | 44 | 68 ± 15 | 3 (13) | 10 (44) | 6 (26) | 6.1 (5.1) | 7 |
|  |  | Non-severe | 85 | 45 | 47 ± 16 | 4 (7) | 6 (10) | 2 (3) | 2.46 (2.3) |  |
| Guner R et al | Turkey | SARI/Critical | 50 | 66 | 62 ± 12 | 10 (20) | 16 (32) | 20 (40) | 5.6 (1.5-38)# | 6 |
|  |  | Mild/pneumonia | 172 | 58 | 48 ± 16 | 20 (12) | 36 (21) | 36 (21) | 2.5 (0.4-28)# |  |
| Song CY et al | China | Severe | 42 | 71 | 56 (48-64) | 4 (10) | 22 (52) | 4 (10) | 8.2 (3.9-19.2) | 6 |
|  |  | Non-severe | 31 | 52 | 48 (37-59) | 2 (7) | 4 (13) | 1 (3) | 3.0 (1.9-5.5) |  |
| Liu J et al | China | Severe | 79 | 58 | 65 (54-71) | 13 (17) | 37 (47) | 2 (3) | 8.83 (4.20-15.53) | 7 |
|  |  | Common | 43 | 61 | 55 (38-66) | 2 (5) | 13 (30) | 0 (0) | 3.11 (1.96-5.00) |  |
| Ma Y et al | China | Severe | 63 | 46 | 53 ± 13 | 6 (10) | 15 (24) | NR | 9.38 ± 10.52 | 6 |
|  |  | Ordinary (Moderate) | 486 | 54 | 45 ± 15 | 23 (5) | 75 (15) | NR | 3.58 ± 3.07 |  |
|  |  | Mild | 86 | 43 | 39 ± 18 | 6 (7) | 15 (17) | NR | 2.73 ± 2.28 |  |
| Chen C et al | China | Critical | 23 | 65 | 68 (63-79) | 9 (39) | 19 (83) | 7 (30) | 7.08 (3.48-12.89) | 8 |
|  |  | Mild | 109 | 56 | 62 (53-70) | 36 (27) | 71 (65) | 24 (22) | 4.10 (2.19-7.51) |  |
| Wang J et al | China | Severe | 8 | 49 | 45 (25-61) | 5 (9) | 13 (24) | 1 (2) | 2.4 (1.4-16.2) | 5 |
|  |  | Moderate | 25 |  |  |  |  |  | 2.3 (1.7-2.9) |  |
|  |  | Mild | 22 |  |  |  |  |  | 1.8 (0.9-2.8) |  |
| CVD = Cardiovascular Disease; DM = Diabetes Mellitus; HT = Hypertension; IQR = Interquartile Range; NLR = Neutrophil-to-lymphocyte ratio; NOS = Newcastle-Ottawa Scale; SD = Standard Deviation; # = min and max data |  |  |  |  |  |  |  |  |  |  |

### Neutrophil-to-lymphocyte ratio (NLR) and severity of COVID-19

There was a total of 5699 patients from a total of 25 included articles comparing on-admission NLR levels in COVID-19 patients with different severity levels. Overall, 1805 patients (32%) had severe COVID-19, and seven studies reported significantly higher proportions of males in the severe COVID-19 group. Compared to the non-severe group, patients with severe COVID-19 were generally older and had more comorbidities, such as diabetes mellitus, hypertension, and cardiovascular diseases. All studies reported higher NLR values on admission in severe COVID-19 patients than non-severe COVID-19 patients (**Table 1, Table S3**). Four out of 25 studies (16%) performed a ROC analysis to determine the optimal cut-off value for NLR value to predict severity^13,16,42,45^. The optimal cut-off value, the area under the curve (AUC), sensitivity, and specificity from the four studies are presented in **Table 3**.

**Table 2.**
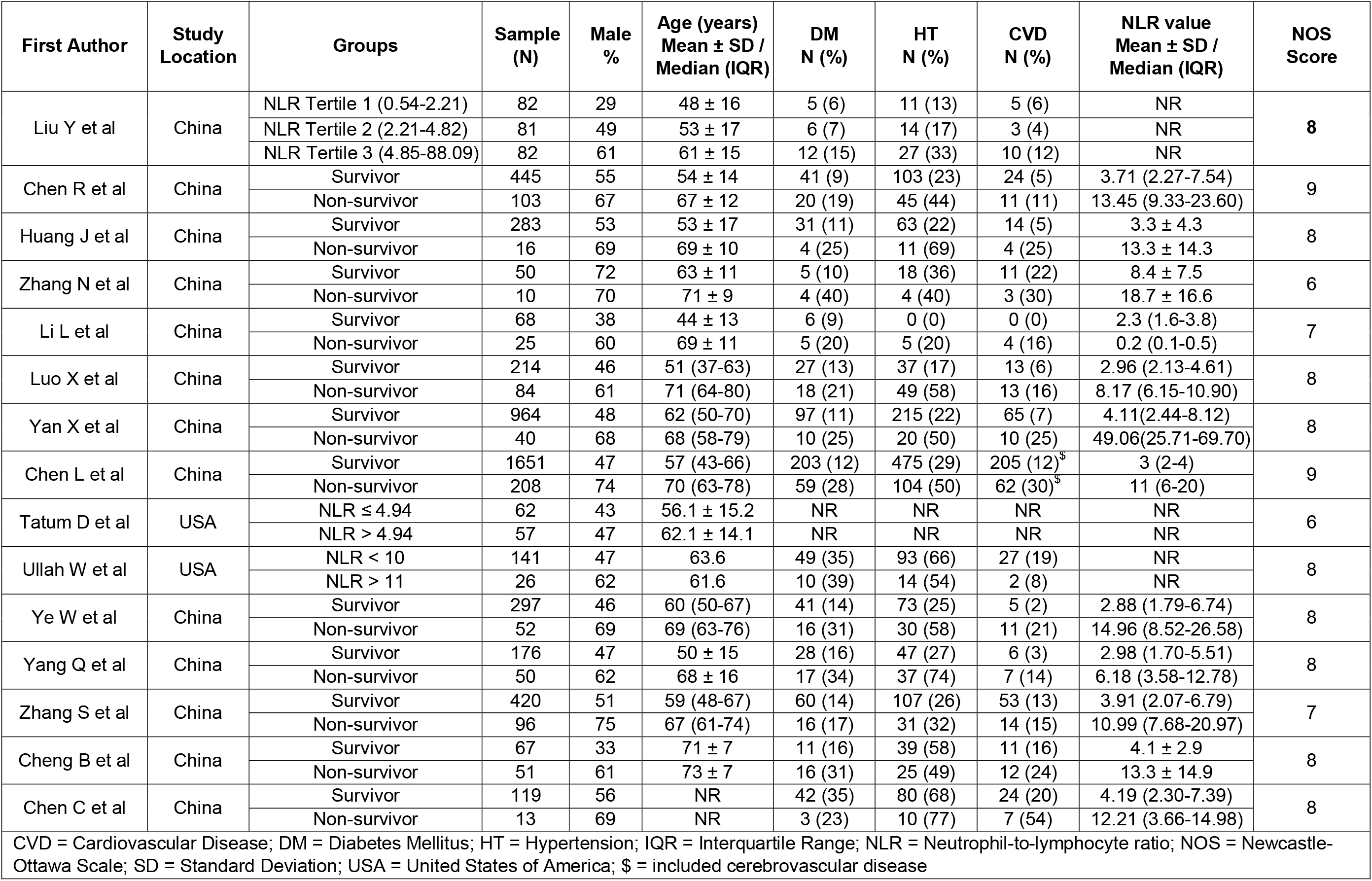
Baseline characteristics of included studies comparing survivors and non-survivors of COVID-19 patients.

**Table 3.** Studies performing Receiver Operating Curve (ROC) analysis.

| First Author | Outcome | Optimal NLR Cut-off Value | Area under the curve (AUC) | Sensitivity (%) | Specificity (%) |
| --- | --- | --- | --- | --- | --- |
| Yang AP et al | Severity | 3.3 | 0.841 | 88 | 63.6 |
| Sun S et al | Severity | 4.5 | NR | 74.07 | 89.89 |
| Song CY et al | Severity | 5.87 | 0.72 | 64 | 81 |
| Ma Y et al | Severity | 4.06 | 0.727 | 61.9 | 76.2 |
| Yan X et al | Mortality | 11.75 | 0.945 | 97.5 | 78.1 |
| Cheng B et al | Mortality | 7.945 | 0.827 | 65.3 | 90.6 |
| NR = Not reported |  |  |  |  |  |

We performed a meta-analysis from 20 eligible articles comparing on-admission NLR levels in COVID-19 patients with different severity groups. From a total of 3859 patients, 1250 patients (32%) experienced severe COVID-19. The pooled analysis showed that severe COVID-19 patients had higher NLR on admission than non-severe patients, with an SMD of 0.88 (95%CI 0.72-1.04) (**Figure 2A**). However, the included studies were significantly heterogeneous (p<0.00001; I^2^=75%). The funnel plot showed that qualitatively, there was no publication bias found in the included studies, and quantitatively Egger’s test showed a low risk of publication bias (p=0.2182) (**Figure 2B**). Sensitivity analysis resulted in no significant changes to the outcome of the analysis. Furthermore, restricting the analysis to only peer-reviewed studies and studies with low risks of bias showed similar pooled results (SMD 0.91; 95%CI 0.73-1.10; I^2^= 79% and 0.87; 95%CI 0.77-0.96; I^2^=46%, respectively) (**Figure S1, S2**). Meta-regression analysis showed that the association between NLR values on admission and severity in COVID-19 patients was not influenced by age (p=0.236), gender (p=0.895), cardiovascular diseases (p=0.886), diabetes mellitus (p=0.880), or hypertension (p=0.222) (**Figure 3A, Figure S3A-D**).

**Figure 2.**
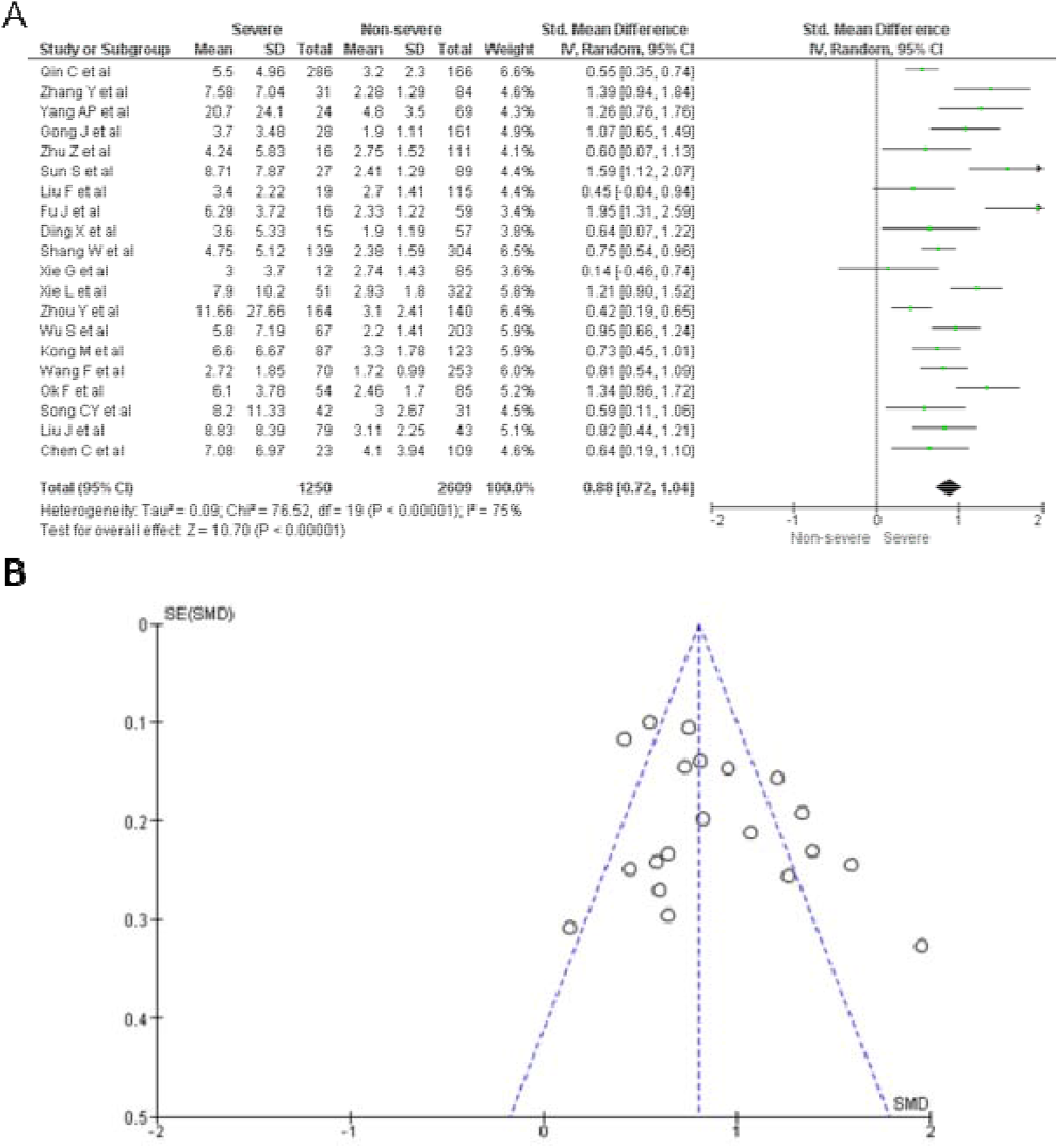
Neutrophil-to-lymphocyte ratio (NLR) value on admission in severe vs. non-severe COVID-19 patients. **A)** Forest Plot for all included studies using the inverse variance random-effect models showing elevated NLR values on admission in severe compared to non-severe COVID-19. **B)** Publication bias analysis of all included studies using the Funnel Plot showing no risk of bias.

**Figure 3.**
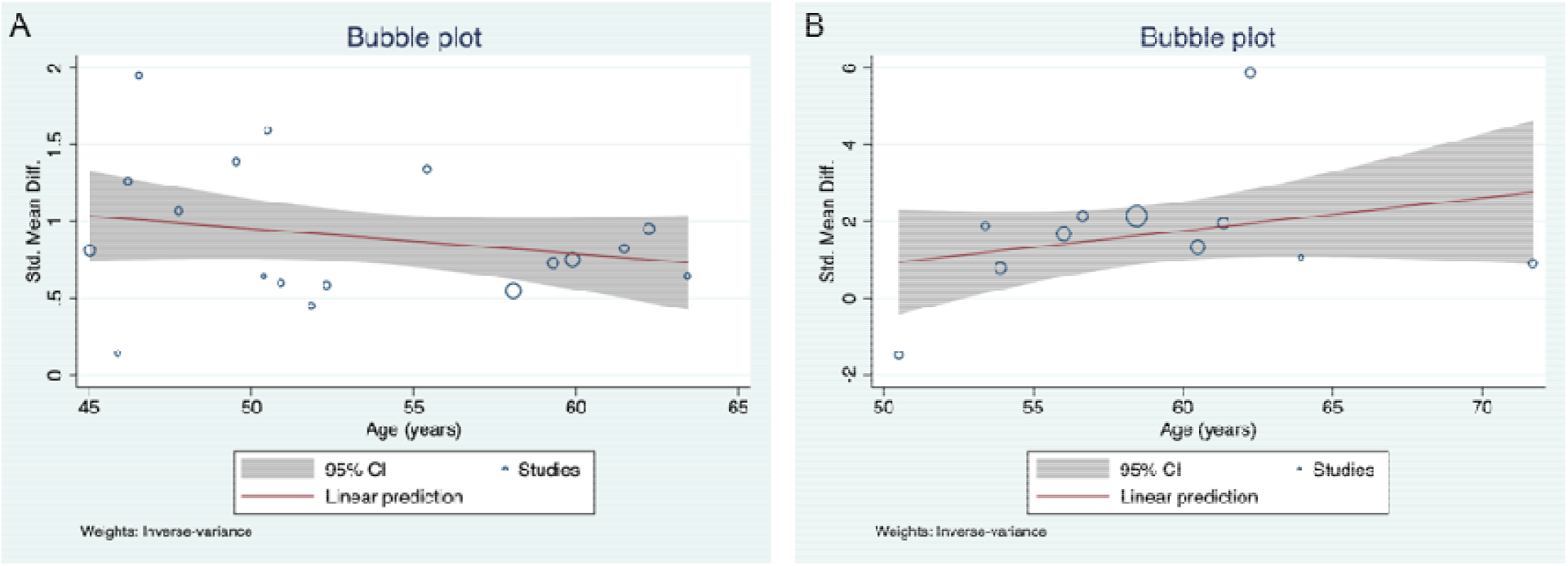
Bubble plot for meta-regression. The association between NLR values on admission and severity of COVID-19 **(A)** and COVID-19 mortality **(B)** was not affected by age (p=0.236; p=0.213, respectively).

### Neutmphil-to-lymphocyte ratio (NLR) and mortality in COVID-19 patients

From a total of 15 studies incorporating 6033 patients with NLR levels on admission in survivor and non-survivor of COVID-19 patients, 822 (14%) died in the hospital. Three of the studies (20%) reported the all-cause mortality of COVID-19 patients in dichotomized NLR values with varying cut-off values (**Table 2**). Generally, those with COVID-19 who died were mostly males, significantly older and had higher proportions of diabetes mellitus, hypertension, and cardiovascular diseases. All studies reported an elevated NLR level on admission in non-survivors compared to survivors except for Li L *et* al.^23^.

Two studies performed a ROC analysis to determine the optimal cut-off value of 11.75 with an AUC value of 0.945 (95%CI 0.917-0.973), a sensitivity of 97.5%, and a specificity of 78.1%^26^; and a cut-off value of 7.945 with an AUC value of 0.827 (95%CI not reported), a sensitivity of 65.3%, and a specificity of 90.6%^44^ (**Table 3**). The multivariable regression comparing patients with low (<11.75) and high (>11.75) NLR levels in Yan X *et al*. resulted in an adjusted odds ratio (AOR) of 44.351 (95%CI 4.627-425.088)^26^.

A meta-analysis was done on 12 eligible studies. From a total of 5502 patients, 748 (14%) were non-survivors. The meta-analysis showed that non-survivors had higher NLR on admission than survivors, with a pooled SMD of 1.68 (95%CI 0.98-2.39). Significant heterogeneity was found among the studies (p<0.00001; I^2^=99%) (**Figure 4A**). The funnel plot showed no publication bias among the included studies, and the calculated Egger’s test showed a low risk of bias (p=0.5315) (**Figure 4B**). Sensitivity analysis by removing one study at a time did not significantly alter the conclusion of the result. Restricting the analysis to only peer-reviewed studies and studies with low risks of bias also showed similar pooled SMD of 1.75 (95%CI 0.95-2.55; I2=99%) and 1.74 (95%CI 1.00-2.48; I2=98%) (**Figure S4, S5**). Furthermore, the association between NLR and mortality in COVID-19 was also unaffected by age (p=0.213), gender (p=0.848), cardiovascular diseases (p=0.889), diabetes mellitus (p=0.526), hypertension (p=0.710) (**Figure 3B, Figure S6A-D**).

**Figure 4.**
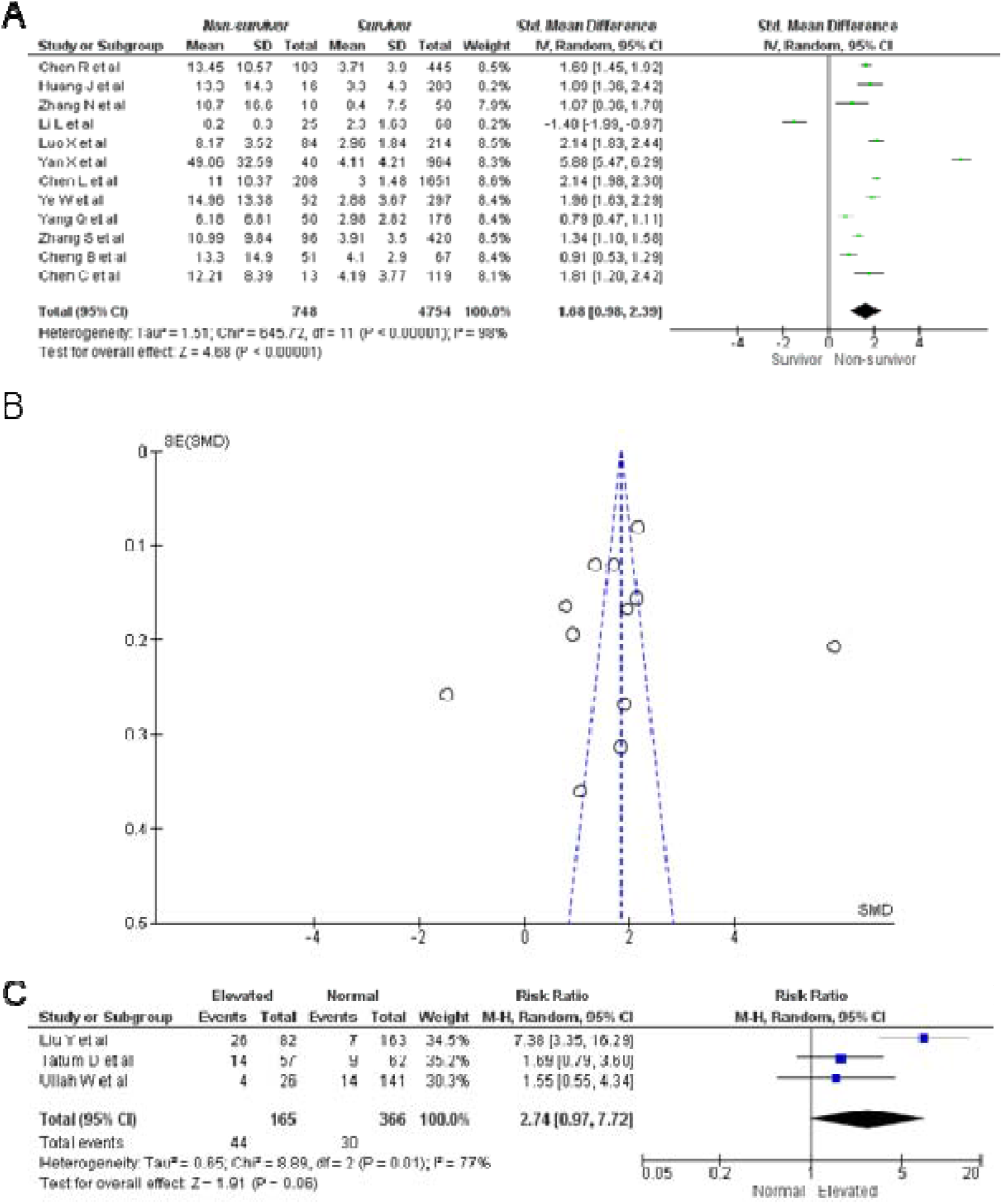
Neutrophil-to-lymphocyte ratio (NLR) value on admission in non-survivor vs. survivor of COVID-19 patients. **A)** Forest Plot for all included studies using the inverse variance random-effects model showing elevated NLR values on admission in non-survivors compared to survivors of COVID-19. **B)** Publication bias analysis of all included studies using the Funnel Plot showing no risk of bias. **C)** Forest Plot using the Mantel-Haenszel random-effects model showing the association between NLR value on admission and all-cause mortality risk.

For the three studies with dichotomized NLR values on admission, a meta-analysis showed that patients with elevated NLR had a higher risk of mortality than those with normal NLR (RR 2.75; 95%CI 0.97-7.72; p=0.06), regardless of the NLR cut-off values used. Significant heterogeneity between the studies was found (p=0.01; I^2^=77%) (**Figure 4C**).

## Discussion

With an increasing number of COVID-19 cases and the limited healthcare capacity, early prediction of COVID-19 severity and mortality is crucial in the patient triage process. Scoring systems such as the Acute Physiology and Chronic Health Evaluation (APACHE) II score was suggested to be a useful clinical tool to predict in-hospital mortality in COVID-19 patients^48^. Other clinical risk scores, such as the COVID-GRAM, are being developed to precisely predict disease progression^49^. However, both scoring systems cannot be implemented in resource-limited healthcare settings, especially in the acute phase, as they heavily rely on advanced laboratory examinations such as serum electrolytes and arterial pH in the APACHE II and lactate dehydrogenase in the COVID-GRAM. Therefore, simpler tools for predicting the severity and mortality of COVID-19 patients in the early stages are urgently needed.

In this study, we performed a systematic review of 38 studies to evaluate the role of NLR levels on admission in predicting the severity and mortality of COVID-19. Our meta-analysis showed that higher NLR values on admission were associated with higher risks of severity and mortality in COVID-19 patients, suggesting that this readily available biomarker can be used to predict the prognosis of COVID-19 patients. However, the differences in NLR values on admission between the survivor and non-survivor patients were greater than those of severe and non-severe patients. Those with high NLR levels on admission had roughly three times the risk of death compared to those with normal NLR levels. Both relationships were shown to be independent of age, gender, diabetes mellitus, hypertension, and cardiovascular diseases.

COVID-19 severity is primarily affected by the innate inflammatory response of the body, where more severe cases were attributed to cytokine storm, a condition where there is an excessive immune response^50^. NLR is a known indicator of systemic inflammation that has been widely used for many conditions, such as predicting in-hospital mortality in sepsis patients, outcomes in cardiovascular diseases, and poor prognosis and higher ICU admissions in acute pancreatitis^51-54^. The biological mechanism underlying this association is that high NLR indicates an imbalance in the inflammatory response, which resulted from increased neutrophil and decreased lymphocyte counts. Inflammatory factors related to viral infection, such as interleukin-6, interleukin-8, and granulocyte colony-stimulating factor, could stimulate neutrophil production^13^. In contrast, systemic inflammation accelerates lymphocyte apoptosis, depresses cellular immunity, decreases CD4+, and increases CD8+ suppressor T-lymphocytes^55,56^.

Compared with other laboratory parameters that predict the prognosis of COVID-19, such as interleukin-6, D-dimer levels, C-reactive protein, or erythrocyte sedimentation rate; NLR is more practical for clinical application as it is easily obtained in routine blood tests^57,58^. Due to the low cost and no need for specific assay equipment, NLR remains a simple, accessible, near real-time, and cost-effective biomarker, especially for healthcare facilities with limited medical resources^59^.

However, to date, no NLR consensus cut-off value has been established to determine normal and elevated NLR values, especially for COVID-19. In determining the optimal cut-off value of NLR, four studies used various NLR values ranging from 3.3 to 5.9 to predict severity^13,16,42,45^, whereas two studies used 7.9 and 11.8 to predict mortality^26,44^. This wide variation indicates that absolute NLR values measured in different populations are hardly comparable and that optimal cut-off values may vary from one population to another.

NLR values were previously reported to vary with age and sex; thus, NLR must be interpreted carefully^60^. Studies have also reported NLR to be race-specific, where different average NLR values were found in different populations^61,62^. In a Chinese population, the reference range of NLR in normal males and females, from a total of 5000 people, was 0.43-2.75 and 0.37-2.87, respectively^60^. The included studies in this meta-analysis generally showed significant differences in age and gender between groups; thus, they could theoretically explain the NLR differences between groups. However, the meta-regression analysis showed that the associations between NLR and COVID-19 severity and mortality were independent of age, gender, and underlying diseases. Therefore, determining the cut-off value is essential for NLR to be used in clinical settings, allowing early risk stratification upon hospital admission.

Our meta-analysis showed significant heterogeneity among the studies. The sensitivity analysis could not determine the source of heterogeneity except for the association between on-admission NLR and severity, where restricting studies to only those with low risk of bias eliminated the heterogeneity. However, in overall, the pooled estimate results were not significantly altered even after performing sensitivity analysis. The identification of heterogeneity among studies with mortality outcomes was not possible due to the possibility of higher variability in treatment protocols among studies with mortality outcomes compared to severity.

To date, our study is the first meta-analysis to describe the predictive values of NLR on admission for the severity and mortality of COVID-19 patients. Additionally, our results showed definitive results that can be directly applied to clinical practice. Moreover, our analysis has emphasized that the association between NLR levels on admission and poor outcomes for COVID-19 was independent of predictors, such as age, hypertension, diabetes mellitus, and cardiovascular diseases.

There are some limitations to this meta-analysis. First, we acknowledge that most of the included studies were primarily conducted in China. Thus, our data might have less clinical relevance in other countries, especially in countries with higher cases and death rates, such as in the United States of America and Europe. Second, we included preprints in the meta-analysis. However, the preprints included had low risks of bias, and further sensitivity analysis to only peer-reviewed studies showed similar results to when preprints were included. Lastly, the studies included in our meta-analysis were all retrospective, except for one, which was a prospective cohort study.14Our meta-analysis demonstrated that NLR on admission is predictive of the severity and mortality in COVID-19 patients, where higher NLR levels are associated with poor outcomes. To date, no optimal cut-off value has been validated across different populations. Therefore, prior to clinical use, further studies should be developed to obtain an exact consensus cut-off value with the optimal sensitivity and specificity. However, our findings support the use of NLR levels to perform early risk stratification in clinical settings, thus allowing patients with higher NLR to be prioritized for healthcare resource allocation.

## Data Availability

All data generated or analyzed during this study are included in the published article and its supplementary information files. The corresponding author (DMS) can be contacted for more information.

## Acknowledgement

None.

## Authors’ Contributions

**DMS:** Idea formulation, article draft writing, data collection and analysis, interpretation of the data, and critical review of the writing; **JC:** data collection and analysis, interpretation of the data, and critical review of the writing; **ADW:** article draft writing, data collection and analysis, and critical review of the writing; **NAAI:** article draft writing, data collection and analysis, and critical review of the writing. All authors have substantially contributed to the formulation of this manuscript. We declare no conflicts of interest.

## References

1. Liu Y-C, Kuo R-L, Shih S-R. COVID-19: The first documented coronavirus pandemic in history. Biomedical Journal. 2020.

2. Sun P, Lu X, Xu C, Sun W, Pan B. Understanding of COVID-19 based on current evidence. Journal of Medical Virology. 2020;92(6):548–551.

3. Chen Y, Liu Q, Guo D. Emerging coronaviruses: Genome structure, replication, and pathogenesis. J Med Virol. 2020;92(4):418–423.

4. Cascella M, Rajnik M, Cuomo A, Dulebohn SC, Di Napoli R. Features, Evaluation and Treatment Coronavirus (COVID-19). StatPearls. Treasure Island (FL); 2020.

5. Terpos E, Ntanasis-Stathopoulos I, Elalamy I, et al. Hematological findings and complications of COVID-19. American Journal of Hematology. 2020;95(7):834–847.

6. Emanuel EJ, Persad G, Upshur R, et al. Fair Allocation of Scarce Medical Resources in the Time of Covid-19. New England Journal of Medicine. 2020;382(21):2049–2055.

7. Lagunas-Rangel FA. Neutrophil-to-lymphocyte ratio and lymphocyte-to-C-reactive protein ratio in patients with severe coronavirus disease 2019 (COVID-19): A meta-analysis. Journal of Medical Virology. 2020;n/a(n/a).

8. Liu Y, Du X, Chen J, et al. Neutrophil-to-lymphocyte ratio as an independent risk factor for mortality in hospitalized patients with COVID-19. The Journal of infection. 2020;81(1):e6-e12.

9. World Health Organization. Report of the WHO-China Joint Mission on Coronavirus Disease 2019 (COVID-19); 2020.

10. Hozo SP, Djulbegovic B, Hozo I. Estimating the mean and variance from the median, range, and the size of a sample. BMC Medical Research Methodology. 2005;5(1):13.

11. Qin C, Zhou L, Hu Z, et al. Dysregulation of immune response in patients with COVID-19 in Wuhan, China. Clinical infectious diseases: an official publication of the Infectious Diseases Society of America. 2020.

12. Zhang Y, Zheng L, Liu L, Zhao M, Xiao J, Zhao Q. Liver impairment in COVID-19 patients: A retrospective analysis of 115 cases from a single centre in Wuhan city, China. Liver international: official journal of the International Association for the Study of the Liver. 2020.

13. Yang A-P, Liu J-P, Tao W-Q, Li H-M. The diagnostic and predictive role of NLR, d-NLR and PLR in COVID-19 patients. International immunopharmacology. 2020;84:106504.

14. Gong J, Ou J, Qiu X, et al. A Tool for Early Prediction of Severe Coronavirus Disease 2019 (COVID-19): A Multicenter Study Using the Risk Nomogram in Wuhan and Guangdong, China. Clin Infect Dis. 2020;71(15):833–840.

15. Zhu Z, Cai T, Fan L, et al. Clinical value of immune-inflammatory parameters to assess the severity of coronavirus disease 2019. Int J Infect Dis. 2020;95:332-339.

16. Sun S, Cai X, Wang H, et al. Abnormalities of peripheral blood system in patients with COVID-19 in Wenzhou, China. Clinica chimica acta; international journal of clinical chemistry. 2020;507:174-180.

17. Liu F, Zhang Q, Huang C, et al. CT quantification of pneumonia lesions in early days predicts progression to severe illness in a cohort of COVID-19 patients. Theranostics. 2020;10(12):5613–5622.

18. Fu J, Kong J, Wang W, et al. The clinical implication of dynamic neutrophil to lymphocyte ratio and D-dimer in COVID-19: A retrospective study in Suzhou China. Thrombosis research. 2020;192:3-8.

19. Ding X, Yu Y, Lu B, et al. Dynamic profile and clinical implications of hematological parameters in hospitalized patients with coronavirus disease 2019. Clinical chemistry and laboratory medicine. 2020;58(8):1365–1371.

20. Chen R, Sang L, Jiang M, et al. Longitudinal hematologic and immunologic variations associated with the progression of COVID-19 patients in China. Journal of Allergy and Clinical Immunology. 2020.

21. Huang J, Cheng A, Kumar R, et al. Hypoalbuminemia predicts the outcome of COVID-19 independent of age and co-morbidity. Journal of medical virology. 2020.

22. Zhang N, Xu X, Zhou LY, et al. Clinical characteristics and chest CT imaging features of critically ill COVID-19 patients. Eur Radiol. 2020:1-10.

23. Li L, Yang L, Gui S, et al. Association of clinical and radiographic findings with the outcomes of 93 patients with COVID-19 in Wuhan, China. Theranostics. 2020;10(14):6113–6121.

24. Shang W, Dong J, Ren Y, et al. The value of clinical parameters in predicting the severity of COVID-19. Journal of medical virology. 2020.

25. Luo X, Zhou W, Yan X, et al. Prognostic value of C-reactive protein in patients with COVID-19. Clinical infectious diseases: an official publication of the Infectious Diseases Society of America. 2020.

26. Yan X, Li F, Wang X, et al. Neutrophil to lymphocyte ratio as prognostic and predictive factor in patients with coronavirus disease 2019: A retrospective cross-sectional study. Journal of medical virology. 2020.

27. Xie G, Ding F, Han L, Yin D, Lu H, Zhang M. The role of peripheral blood eosinophil counts in COVID-19 patients. Allergy. 2020;n/a(n/a).

28. Xie L, Wu Q, Lin Q, et al. Dysfunction of adaptive immunity is related to severity of COVID-19: a retrospective study. Therapeutic Advances in Respiratory Disease. 2020;14:1753466620942129.

29. Chen L, Yu J, He W, et al. Risk factors for death in 1859 subjects with COVID-19. Leukemia. 2020.

30. Zhou Y, Guo S, He Y, et al. COVID-19 Is Distinct From SARS-CoV-2-Negative Community-Acquired Pneumonia. Frontiers in Cellular and Infection Microbiology. 2020;10(322).

31. Wu S, Du Z, Shen S, et al. Identification and validation of a novel clinical signature to predict the prognosis in confirmed COVID-19 patients. Clinical Infectious Diseases. 2020.

32. Tatum D, Taghavi S, Houghton A, Stover J, Toraih E, Duchesne J. Neutrophil-to-Lymphocyte Ratio and Outcomes in Louisiana Covid-19 Patients. Shock (Augusta, Ga). 2020.

33. Ullah W, Basyal B, Tariq S, et al. Lymphocyte-to-C-Reactive Protein Ratio: A Novel Predictor of Adverse Outcomes in COVID-19. Journal of clinical medicine research. 2020;12(7):415–422.

34. Kong M, Zhang H, Cao X, Mao X, Lu Z. Higher level of neutrophil-to-lymphocyte is associated with severe COVID-19. Epidemiology and infection. 2020;148:e139.

35. Wang F, Qu M, Zhou X, et al. The timeline and risk factors of clinical progression of COVID-19 in Shenzhen, China. Journal of Translational Medicine. 2020;18(1):270.

36. Ye W, Chen G, Li X, et al. Dynamic changes of D-dimer and neutrophil-lymphocyte count ratio as prognostic biomarkers in COVID-19. Respiratory Research. 2020;21(1):169.

37. Yang Q, Zhou Y, Wang X, et al. Effect of hypertension on outcomes of adult inpatients with COVID-19 in Wuhan, China: a propensity score-matching analysis. Respiratory Research. 2020;21(1):172.

38. Liao D, Zhou F, Luo L, et al. Haematological characteristics and risk factors in the classification and prognosis evaluation of COVID-19: a retrospective cohort study. The Lancet Haematology. 2020;7(9):e671-e678.

39. Ok F, Erdogan O, Durmus E, Carkci S, Canik A. Predictive values of blood urea nitrogen/creatinine ratio and other routine blood parameters on disease severity and survival of COVID-19 patients. Journal of medical virology. 2020.

40. Guner R, Hasanoglu I, Kayaaslan B, et al. COVID-19 experience of the major pandemic response center in the capital: Results of the pandemic’s first month in Turkey. Turkish journal of medical sciences. 2020.

41. Zhang S, Guo M, Duan L, et al. Development and validation of a risk factor-based system to predict short-term survival in adult hospitalized patients with COVID-19: a multicenter, retrospective, cohort study. Critical care (London, England). 2020;24(1):438.

42. Song C-Y, Xu J, He J-Q, Lu Y-Q. COVID-19 early warning score: a multi-parameter screening tool to identify highly suspected patients. *medRxiv*. 2020:2020.2003.2005.20031906.

43. Liu J, Liu Y, Xiang P, et al. Neutrophil-to-lymphocyte ratio predicts critical illness patients with 2019 coronavirus disease in the early stage. Journal of Translational Medicine. 2020;18(1):206.

44. Cheng BG, Tang-Meng Tong Y, Chen J, Huang L, Zhoue J. Clinical Features Predicting Mortality Risk in Older Patients with COVID-19. *Available at SSRN 3569846*. 2020.

45. Ma YS, Nannan Fan Y, Wang J, et al. Predictive Value of the Neutrophil-to-Lymphocyte Ratio (NLR) for Diagnosis and Worse Clinical Course of the COVID-19: Findings from Ten Provinces in China. *Available at SSRN*. 2020.

46. Chen C, Yi ZJ, Chang L, et al. The characteristics and death risk factors of 132 COVID-19 pneumonia patients with comorbidities: a retrospective single center analysis in Wuhan, China. *medRxiv*. 2020:2020.2005.2007.20092882.

47. Wang J, Li Q, Yin Y, et al. Neutrophilia and Lymphopenia Occur Coincidentally with Pneumonia Progression in Severe COVID-19 Patients. *Available at SSRN 3582712*. 2020.

48. Zou X, Li S, Fang M, et al. Acute Physiology and Chronic Health Evaluation II Score as a Predictor of Hospital Mortality in Patients of Coronavirus Disease 2019. Critical care medicine. 2020;48(8):e657-e665.

49. Liang W, Liang H, Ou L, et al. Development and Validation of a Clinical Risk Score to Predict the Occurrence of Critical Illness in Hospitalized Patients With COVID-19. JAMA internal medicine. 2020.

50. Coperchini F, Chiovato L, Croce L, Magri F, Rotondi M. The cytokine storm in COVID-19: An overview of the involvement of the chemokine/chemokine-receptor system. Cytokine & Growth Factor Reviews. 2020;53:25-32.

51. Sari R, Karakurt Z, Ay M, et al. Neutrophil to lymphocyte ratio as a predictor of treatment response and mortality in septic shock patients in the intensive care unit. Turkish journal of medical sciences. 2019;49(5):1336–1349.

52. Bhat T, Teli S, Rijal J, et al. Neutrophil to lymphocyte ratio and cardiovascular diseases: a review. Expert Rev Cardiovasc Ther. 2013;ll(l):55-59.

53. Azab B, Jaglall N, Atallah JP, et al. Neutrophil-lymphocyte ratio as a predictor of adverse outcomes of acute pancreatitis. Pancreatology. 2011;ll(4):445-452.

54. Wang X, Zhang G, Jiang X, Zhu H, Lu Z, Xu L. Neutrophil to lymphocyte ratio in relation to risk of all-cause mortality and cardiovascular events among patients undergoing angiography or cardiac revascularization: A meta-analysis of observational studies. Atherosclerosis. 2014;234(1):206–213.

55. Menges T, Engel J, Welters I, et al. Changes in blood lymphocyte populations after multiple trauma: association with posttraumatic complications. Crit Care Med. 1999;27(4):733–740.

56. Channappanavar R, Perlman S. Pathogenic human coronavirus infections: causes and consequences of cytokine storm and immunopathology. Semin Immunopathol. 2017;39(5):529–539.

57. Ponti G, Maccaferri M, Ruini C, Tomasi A, Ozben T. Biomarkers associated with COVID-19 disease progression. Critical Reviews in Clinical Laboratory Sciences. 2020:1-11.

58. Simadibrata DM, Lubis AM. D-dimer levels on admission and all-cause mortality risk in COVID-19 patients: a meta-analysis. Epidemiology and Infection. 2020:1-24.

59. Guo J, Fang J, Huang X, et al. Prognostic role of neutrophil to lymphocyte ratio and platelet to lymphocyte ratio in prostate cancer: A meta-analysis of results from multivariate analysis. Int J Surg. 2018;60:216-223.

60. Wu L, Zou S, Wang C, Tan X, Yu M. Neutrophil-to-lymphocyte and platelet-to-lymphocyte ratio in Chinese Han population from Chaoshan region in South China. BMC cardiovascular disorders. 2019;19(1):125–125.

61. Azab B, Camacho-Rivera M, Taioli E. Average values and racial differences of neutrophil lymphocyte ratio among a nationally representative sample of United States subjects. PLoS One. 2014;9(11):e112361.

62. Lee JS, Kim NY, Na SH, Youn YH, Shin CS. Reference values of neutrophil-lymphocyte ratio, lymphocyte-monocyte ratio, platelet-lymphocyte ratio, and mean platelet volume in healthy adults in South Korea. Medicine (Baltimore). 2018;97(26):e11138.

